# Metabolic health tracking using Ultrahuman M1 continuous glucose monitoring platform in non- and pre-diabetic Indians: a multi-armed observational study

**DOI:** 10.1101/2023.09.20.23295642

**Authors:** Monik Chaudhry, Mohit Kumar, Vatsal Singhal, Bhuvan Srinivasan

## Abstract

**Background:** CGM-based tracking is expanding in non-diabetic groups to meet wellness and preventive care needs. However, data is limited on short-term outcomes for glycemic control, insulin resistance and correlation of algorithm-derived score to known glycemic metrics in controlled settings, making benchmarking difficult. This is especially true for the high-risk Indian/South Asian demographic.

**Objectives:** To examine changes resulting from the Ultrahuman (UH) M1 CGM application-with concomitant FitBit tracker use in patterns of glucose variability (GV). Evaluate GV correlations with stress, sleep duration, inflammation, and activity. Examine correlations between UH metabolic score (UH-MS) and biomarkers of dysglycemia and insulin resistance.

**Methods:** Participants (N=53 non-diabetic, 52 pre-diabetic) wore the UH-M1 CGM and FitBit tracker for a 14-day period. HsCRP, cortisol, OGTT, HbA1c, HOMA-IR levels, and standard blood profile measurements were obtained.

**Results:** Mean glucose levels, restricted time in range (70-110mg/dL), and GV metrics were significantly different between non- and pre-diabetics and displayed improvements with M1 use. Strong correlations of specific GV metrics with inflammation were found in pre-diabetics, with modest correlation between sleep and activity in non-diabetics. Elevated HOMA-IR, HbA1c, and OGTT were linked with J-index and high blood glucose index in pre-diabetics, and low blood glucose index in non-diabetics. UH-MS displayed a strong inverse relationship with insulin resistance and glucose dysregulation.

**Conclusions:** The study presents the first guidance values of glycemic indices of non- and pre-diabetic Indians and supports the notion that short-duration CGM use with algorithm scores can affect positive changes in glucose management.

## Introduction

Type 2 diabetes mellitus (T2DM) and its associated metabolic conditions are known global pandemics with an estimated prevalence of 422 million.^1^ Similarly, the burden of prediabetes (intermediate hyperglycemia) is increasing at an aggressive rate with a projected estimate of 8.0 % (454 million) by 2030 and 8.6% (548 million) by 2045.^1,2^ Prediabetes is defined by glycemic variables that are intermediate between healthy and diabetic ranges. Hyperglycemia is known to upregulate chronic inflammatory markers, and cellular stress such as increased reactive oxygen species (ROS) generation which leads to a condition termed insulin resistance, wherein cells become insensitive to insulin and have lower activity-dependent glucose uptake.^3, 4^ These physiological and cellular changes propel the individual towards the diabetic “state”, and it is estimated that approximately 5-10% of pre-diabetics convert to diabetics per year worldwide with a large variation depending on diagnostic criteria and geography^5^.

Interestingly, a considerable proportion of pre-diabetic patients can revert to normoglycemia if proper corrective measures are implemented including consistent tracking of blood glucose levels and complementing lifestyle modification.^6^ The widely accepted American Diabetes Association (ADA) guidelines, therefore, strongly emphasize the adoption of these non-pharmacological management and lifestyle modification techniques as soon as a person is diagnosed as a pre-diabetic.^7^ The extension of these health management measures has reached the wellness sector in recent times, attesting to their real-world effectiveness.

Several risk-scoring diagnostics and biochemical tests (random blood glucose, fasting blood glucose [FBG], 75g-oral glucose tolerance test [OGTT], glycated hemoglobin [HbA1c]) usually in combination, are widely used for pre-diabetes and diabetes surveillance.^8, 9^ In recent times, the practice of continuous glucose monitoring (CGM) using subcutaneous sensors has revolutionized the concept of real-time glucose tracking and offered dependable solutions for screening both diabetic and pre-diabetic individuals.^10, 11^ CGM-mediated real-time tracking of glycemic variables improves the chances of detecting glycemic deviations in basal, night-time, and postprandial conditions, and derive correlations between dietary- and exercise-related changes in lifestyle and glucose tolerance. These advantages have accelerated the adoption of CGMs among high-performance athletes, fitness-oriented healthy individuals, and suspected pre- diabetics.^12,13^ The challenge remains in developing easy-to-understand metrics and user interfaces that promote better adoption and compliance, and enhance the predictive component of interpreted glycemic trends by evidence-based correlation.

The Ultrahuman (UH) M1 platform consists of a CGM sensor, application (app)-based analytics, and timely fitness advice provided by certified experts. ^15^ The captured glucose data is used to generate the daily user-specific metabolic score (MS), which is a holistic snapshot of a user’s daily glucose regulation patterns (see Methods). ^16^ The app also prompts lifestyle changes by providing actionable nudges and alerts to the user (e.g., a prompt to move if the glucose level rises above the target range).

In South Asia, especially India, CGMs are predominantly used for diabetes evaluation.^17^ While wellness and lifestyle monitoring apps enjoy reasonable following, the data include user-uploaded, non-biomarker information such as food logs, step counts, sleep duration, etc., which provide a general overview of health but cannot be substantially correlated to clinical biomarkers. This gap in biomarker-based tracking is especially crucial for India which is increasingly being known as the “Diabetic capital of the world”.^18^ In a recently published, nationwide survey, the overall prevalence of diabetes was calculated to be 11.4%, and for prediabetes 15.3%.^19^ More concerning was that individuals with intermediate fasting glucose (IFG, 100-125mg/dL) had tripled since the last survey in 2017.^20^ Alongside are the findings of the CUREs longitudinal study, which reports that 58.9% of pre-diabetic Indians convert to diabetes over a 10-year period.^21^ Taken together, there is substantial evidence of Indian and South Asians having a high susceptibility to metabolic syndrome and a silent epidemic is most likely underway in this population.

Although the scope of remote monitoring and use of apps has improved after the COVID pandemic, the lack of population-scale, curated digital data for evidence-based profiling of health status creates a gap in usable glycemic benchmarks for Indian and South Asian profiles. This results in a dearth of clinically relevant data to differentiate between the healthy and at-risk populations based on CGM findings, impacting the design of point-of-care, customized lifestyle management interventions, which is the main premise of non-pharmacological management for diabetes prevention prescribed by ADA and other organizations.

Therefore, to address this gap, we undertook a controlled, multi-arm observational study to simultaneously derive glycemic variability (GV) data in non-diabetics (healthy) and pre-diabetics, correlate these with established markers of inflammation, stress, and lifestyle indicators of sleep, step count, and heart rate; examine the relationship between these biomarkers and MS, and finally obtain profiles of glucose tolerance following a 14-day use of the UH-M1 platform for both groups.

## Methods

### Study design and participants

This prospective two-arm parallel-group observational study was conducted across multiple urban diabetes clinics and hospitals (N=9) across the states of Delhi, Karnataka, Telangana, Gujarat, and Tamil Nadu within India. Following this, participants were recruited in an enrolment period that spanned from September 2022 to December 2022.

The overarching aim of this study was to assess CGM-derived GV indices and their correlation with clinical biomarkers in healthy and pre-diabetic individuals to generate reference data on metabolic health for this age and geographic group. The dataset would also be used to a) investigate MS correlation with well-established clinical biomarkers of stress, sleep, inflammation, insulin resistance, and glucose intolerance, and b) form the basis of updating MS. MS is a proprietary algorithmic output that has been developed by Ultrahuman Pvt Ltd., for the purpose of metabolic fitness tracking and management.^16^

Participants (males and females) were included in the study if they were between 25-50 years of age (both inclusive) and had body mass index (BMI) within 20 – 30 kg/m^2^ range. They were required to comply with the advised use of CGM (Abbott FreeStyle Libre^22^, activity tracker (Fitbit Inspire 2), and the UH Application. The exclusion criteria consisted of a history of acute or subacute infection (within the last three months) and chronic illnesses (including T1 (Type 1)- and T2DM, and cardiac disease), anemia, endocrine disorders, and autoimmune conditions. Individuals taking antimicrobials, including antibiotics, antivirals, and antifungals were also ineligible for participation.

The study was conducted in compliance with the International Conference of Harmonization/Good Clinical Practice guidelines (ICH-GCP) and the Declaration of Helsinki. Participants voluntarily signed a written informed consent form prior to participation and were allowed to withdraw from the study at any time. The study protocol was reviewed and approved by the ethics committees of all the participating centers. Ethics clearance was secured individually at each site involving either the hospital’s ethics committee or an independently instituted ethics committee for smaller clinics. The details of the trial site, ethics committee, and date of approval are provided in a tabular format in **Supplementary Table 7**. This study is registered in the Clinical Trials Registry - India CTRI/2022/08/044808.

### Study procedure

Random sampling method was used to recruit eligible subjects. During the screening visit (between -3 to -1 day from the baseline [inclusion] visit), a detailed medical, medication-related, and family history was acquired. Demographic data, anthropometric measurements, and vital signs were recorded and blood samples were obtained to estimate FBG, and glycated HbA1c levels. Potential participants also underwent an OGTT test. Based on the results obtained, the subjects were then screened for eligibility and those selected were divided into two groups: healthy/non-diabetic (FBG: 79-99 mg/dl; HbA1c: 4.0-5.6 % and 2-hour plasma glucose during 75-g OGTT below 140 mg/dL) and pre-diabetics (FBG: 100-125 mg/dl; HbA1c: 5.7-6.4 % and 2-hour plasma glucose during 75-g OGTT: 140–199 mg/dL) based on the ADA criteria of Screening and Diagnostic Tests for Prediabetes.^7^

At day 0 (baseline visit), the eligibility was reconfirmed by repeating the OGTT and a general physical examination. Details regarding the CGM and UH-M1 application were also explained to the participants during this visit. The app was installed on the smartphone of the subject, and he/she was trained on the features of the app and its use. Once the subject was familiar with the app, the CGM was attached to the upper arm (preferably left) and activated followed by the initializing of the app. The participants were asked to follow a regular daily routine and log (food information) the same on the UH-M1 app daily. He/she was instructed to contact the investigator or the team in case of any difficulties while using the app.

Adverse reactions (if any) were planned to be coded using the MedDRA central coding dictionary, version 25. All medications were to be coded using the WHO-DD, September 1, 2019, or later. Preferred ATC coding was planned to be applied to encode medications use.

A second follow-up visit was arranged between days 5-7 of the trial period. The tests conducted on this day included an OGTT and a general physical examination. This OGTT visit was postponed in subjects with any concomitant indigestion, gastric irritation, or vomiting. Data collection ended on day 14 of CGM and app use, followed by a final, physical examination and laboratory investigations. In the case of sensor failure (sensor stopped reporting values or widely fluctuating measurements) the endpoint occurred earlier. This session was termed as the “End of study” (EOS) visit. In addition, the participants also completed a satisfaction feedback form and a Pittsburgh Sleep Quality Index (PSQI) sleep self-assessment questionnaire during this visit. ^23^ Subjects experiencing any temporary health issues, technical difficulties in using CGM, CGM data collection failure, or non-compliance with the app were discontinued/withdrawn from the study.

### Study endpoints

The primary endpoints included CGM-based glucose indices over 14 days period such as the mean glucose levels as described by a 24-hour profile during 2 weeks; time in glucose ranges (TIR: 70-180 mg/dL for “acceptable” diabetes glucose range; TAR: time-above-range >180 mg/dL and TBR: time-below-range <70 mg); GV as measured by the standard deviation (SD) and the coefficient of variation (CV); and the mean amplitude of glycaemic excursions (MAGE), defined as the arithmetic mean of the amplitude of glucose excursions that are greater than the standard deviation of the glucose values. In addition, for the preventive/wellness use case, the UH-CGM application employs a tighter target range of 70- 110mg/dL; and hence it was also computed for the healthy and pre-diabetic groups post-facto after study completion. Daily MS scores were generated for each participant across the study period which were then used for correlation analyses (representative snapshot of MS display on the app interface is depicted in Supplementary Figure S1).

The secondary endpoints included changes in FBG levels from day 0 -15, the correlation between GV indices and sleep duration, step count, heart rate (acquired from fitness tracker use), and blood-based biomarkers such as stress (serum cortisol), inflammation (serum Hs-C reactive protein (Hs-CRP)). Additional samples to catalog gut microbiome, and urine metabolites were also acquired for future analyses and are not within the scope of this manuscript.

### Statistical analysis

Data were analyzed between day 2-14 of CGM use to rule out differences in sensor application across participants and known variability of sensor output in the first 24h of sensor activation^24^. Data were analyzed using the R Software version 4.2.2.^25^ Intent-to-Treat (ITT) set (included all subjects who were enrolled in the study) and Per Protocol (PP) set (included all subjects who completed the study procedures as per the planned protocol) were defined for analyzing the data. Normality tests were performed to select the appropriate test and the outliers were removed following ±3SD for normal distributions, and beyond three times lowest and highest interquartile range (IQR) for non-parametric data. Categorical data were presented as frequencies and proportions and compared using the Chi-square test with Yates correction or Fischer’s exact t-test, as appropriate. Continuous data were presented as mean with SD or median with interquartile range and were compared using unpaired t-test, or Mann-Whitney U test, as appropriate. In addition to summary statistics, the differences in the primary endpoint results between non-diabetic and pre-diabetic subjects’ groups were compared using statistical models. Least-squares means (LSM), visit differences in LSM, and the corresponding 95% confidence intervals (CIs) for the subject group differences were estimated using the model. Receiver operating characteristic (ROC) curve analysis was used to assess the predictive value of CGM-based GV indices in prediabetes. For the secondary endpoints, Pearson correlation or Spearman coefficients were calculated and presented in graph and tabular outputs to assess the association between the clinical biomarkers, and interstitial glucose. Linear models were also used to explore these associations. All statistical tests were conducted at a 2-sided alpha level of 0.05 and a 2-sided 95% CI was provided.

## Results

### Patient disposition and trends

A total of 151 subjects were screened of which 105 met all the inclusion criteria and participated in the study (**Figure 1**). All the 105 enrolled participants completed the study and there were no drop-outs. Both the healthy (n=53) and pre-diabetic (n=52) groups were well-matched with respect to baseline demographics and clinical characteristics (**Supplementary Table S1**). The mean BMI was slightly higher in healthy, but there was no significant difference between the groups. Physical examination and medical history were well within the inclusion parameters. Although there was a higher prevalence of familial diabetes in the pre-diabetic cohort and a higher rate of familial hypertension in the non-diabetic group, the differences were not statistically supported (data not shown). No serious adverse events were reported during the study.

**Figure 1.**
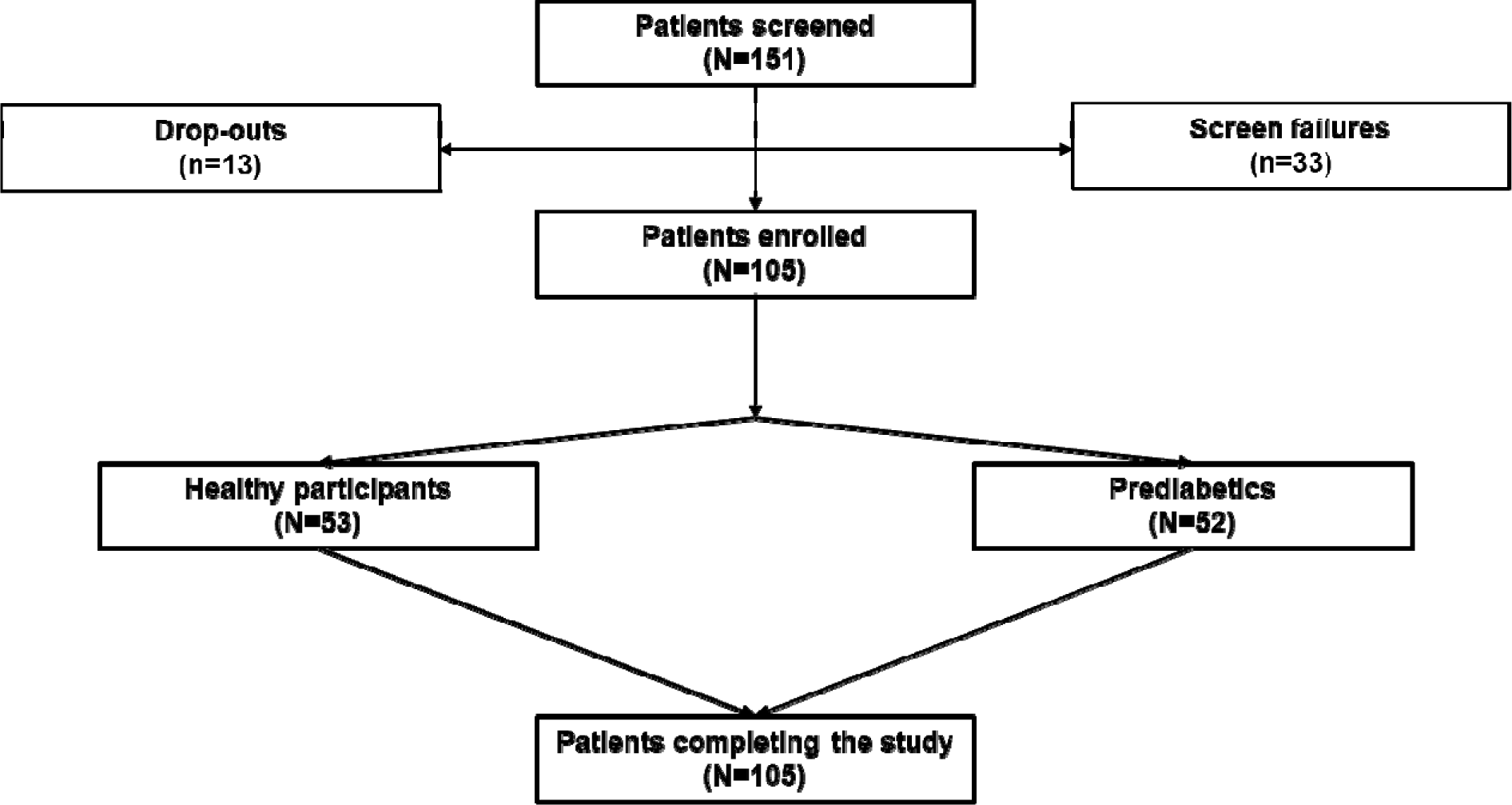
Patient disposition.

### Primary outcomes: Changes in mean blood glucose and TIRs, across non-diabetics and pre-diabetics over time

Over the years, a wealth of studies have contributed to the generation of a set of CGM-derived glycemic indices, which have been used in a variety of ways to assess metabolic health.^26^ Normal reference ranges of some of these markers are available in small studies with diverse ethnic representation; however, information on South Asians derived from controlled studies is scarce.^27^

In our cohort, we observed a significant difference in daily the mean glucose levels detected by UH-M1 between the healthy (Mean ± SD: 102.4±11.78 mg/dL) and pre-diabetic (Mean ± SD: 112.2 ±14.25 mg/dL) individuals and this difference extended over the entire duration of 14 days (Two-way ANOVA, main effect, cohort: p<0.0001; interaction cohort x day: p<0.01; **Figure 2A**). It is noteworthy that there was a significant downward trend over time in mean glucose levels in both groups (main effect, day: p<0.0001). The mean percentage of CGM-based TIR between day 2 to day 14 was better in healthy individuals (95.3 % ±10.43) than in the pre-diabetic group (94.6 % ±9.4), however, there was no statistically significant difference between the two groups (Two-way ANOVA, main effect, cohort: p=0.91; main effect, day: p=0.91, **Figure 2B**).

**Figure 2.**
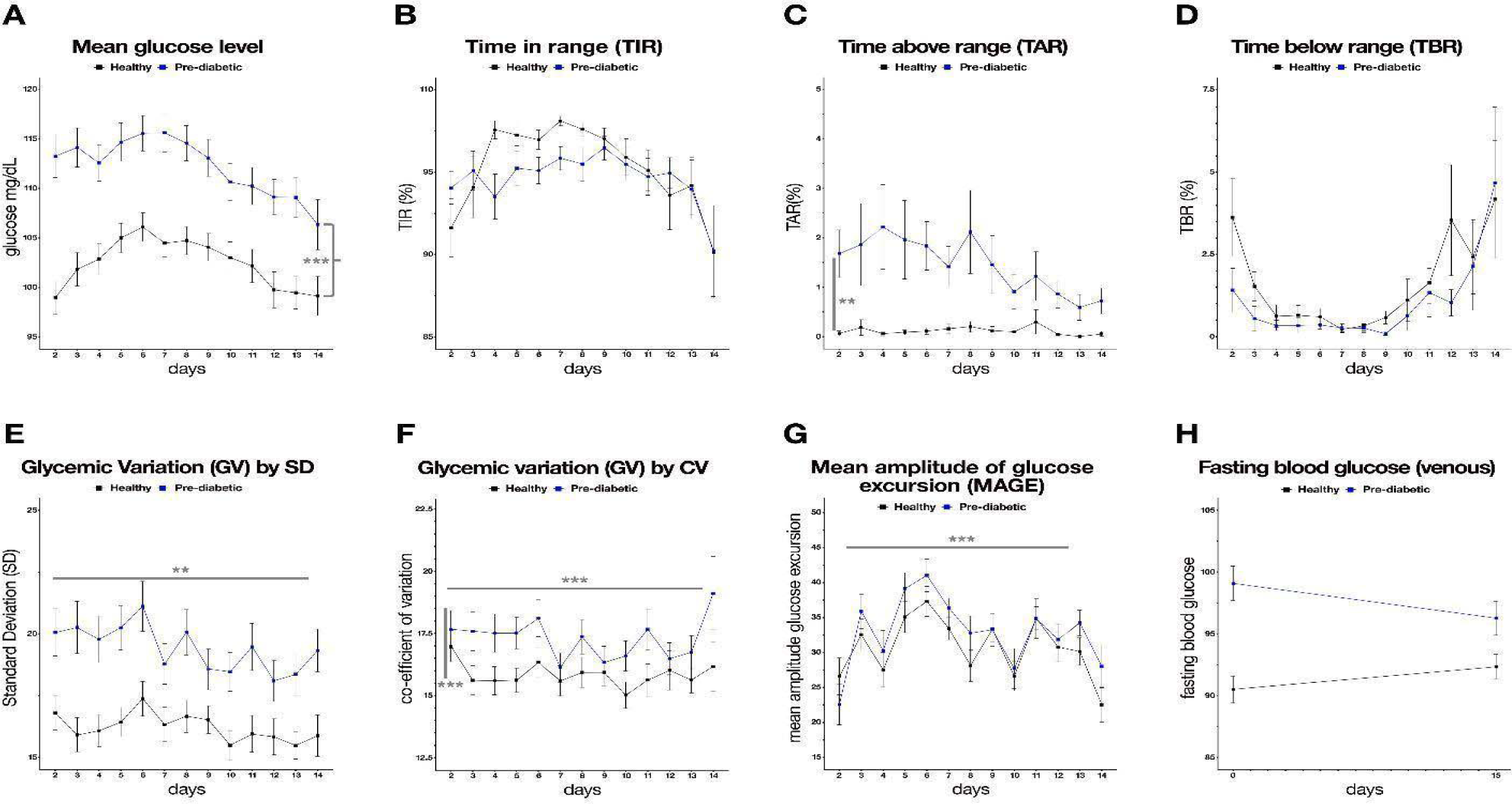
Primary outcome measures in healthy vs. pre-diabetic within the stipulated time-frame. **CV: Coefficient of variation; MAGE: Mean amplitude of glycemic excursion; SD: Standard deviation. *, **, *** denotes p<0.05, 0.01 and 0.001, by Two -way ANOVA (see text for details)**

Furthermore, TIR adherence tapered over the period of 14 days, especially towards the end of the study in healthy and pre-diabetics (**Figure 2B**). The UH-M1 application uses a relatively tighter target range of 70-110mg/dL glucose (as compared to the normal TIR of 72-180 mg/dL). Post-facto calculation for this restricted TIR (rTIR), revealed extremely significant differences between the groups and across days (Two-way ANOVA, main effect, cohort: p<0.0001; main effect, day: p<0.0001, interaction cohort x day: p<0. 0.00001, **Supplementary Figure S2**). Pre-diabetics consistently had lower dwell times in the optimal rTIR range as compared to non-diabetics, and both groups appeared to reach comparable rTIR values by the end of the study period. For TAR (>180 mg/dL glucose), there was a statistically significant difference (Two-way ANOVA, main effect, cohort: p<0.01; main effect, day, ns, interaction cohort x day: ns) between pre-diabetics (mean 1.4% ±4.15) and healthy individuals (mean 0.1% ±0.72); and the healthy participants had negligible hyperglycemic events **(Figure 2C)**. Interestingly, a clear downward trend was detected in pre-diabetics over time (**Figure 2C**). There were no distinct trends in TBR identifying hypoglycemic events (< 72 mg/dL glucose), in healthy (mean 1.6%±6.43) and pre-diabetics (mean 1.0%± 5.99) with a higher level of such events in both groups towards the end of the observation period (**Figure 2D**).

### Primary outcomes: Changes in glycemic variability indices, across non-diabetics and pre-diabetics over time

Next, we measured a variety of CGM-derived indices of cumulative glycemic variability in an effort to identify which of the metrics efficiently differentiated between non-diabetics and pre-diabetics and captured changes in trends due to app-based tracking. Results revealed that GV, as measured by standard deviation (GV by SD), captured significant across-group differences in the analyses period (19.4±6.51 pre-diabetics vs 16.2±4.85 non-diabetics, Two-way ANOVA, main effect, cohort: p<0.001; main effect, day: p<0.001, interaction cohort x day: ns, **Figure 2E**). In comparison, GV as measured by the coefficient of variation (GV by CV), had more overlaps between groups, with milder but significantly different values (17.3± 5.67 pre-diabetics vs 15.8±4.57 non-diabetics, Two-way ANOVA, main effect, cohort: p<0.05; main effect, day: p<0.05, interaction cohort x day: ns; **Figure 2F**). Both GV by SD and GV by CV indices displayed a gradual decrease over the trial period in both groups.

Mean amplitude of glycaemic excursions (MAGE) values showed across-day improvements, but the level of decrease was not as significant as GV, and there was no distinction between healthy (30.7±15.99) and pre-diabetic (32.9±18.19) individuals (Two-way ANOVA, main effect, cohort: p=0.083; main effect, day: p<0.0001, interaction cohort x day: ns; **Figure 2G**).

Finally, FBG levels were monitored at the beginning and end of the study to provide an external anchor point for CGM-derived values. FBG increased modestly from 90.5±7.90 mg/dl at baseline to 92.3±7.28 mg/dl at day 15 in healthy individuals while it decreased from 99.1±10.11 mg/dl at baseline to 96.3±9.84 mg/dl in pre-diabetics **(Figure 2H)**. Given the convergent trend of FBG values, there were no significant differences between groups or across time in each group (ANCOVA model with treatment as fixed effect and FBS values at baseline visit as covariate, p=0.81).

### Secondary outcomes: Correlation of CGM-derived glycemic metrics with biomarkers associated with metabolic syndrome

A wealth of evidence supports a strong relationship between stress-related markers such as inflammation, disturbed sleep, reduced physical exercise, and impaired cortisol and the development of prodromal conditions like prediabetes. ^28-30^ Several data points are also available for the Indian / South Asian demographic group as well.^31^ However, these reports do not utilize CGM-derived metrics. To address this gap, we carried out a correlation analysis for sleep, stress, inflammation, heart rate, and step count in our cohort with the following seven established glycemic indices: J-index, high blood glucose index (HBGI), low blood glucose index (LBGI), average daily risk range (ADRR), MAGE, mean of daily differences (MODD) and continuous overall net glycemic action (CONGA).^32^ Here we highlight the main results with all data presented in **Supplementary Tables S2-S5**.

The strongest correlation was found between GV indices and inflammation as measured by serum Hs-CRP levels (**Supplementary Table 2, Figure 3 A-C**). The J index, HBGI, and LBGI revealed a strong positive correlation in pre-diabetics, but not in non-diabetic individuals. Surprisingly, there was little correlation with stress as measured by serum cortisol levels and any of the GV indices in either group **(Supplementary Table S3)**. This may be because GV was calculated over 2-14 days whereas cortisol levels were measured on day 0 and day 15. Sleep duration was negatively correlated with HBGI in non-diabetic but not pre-diabetic individuals **(Supplementary Table S4, Figure 3D)**. Interestingly, CONGA was negatively correlated with sleep data in both groups, the significance of which is as yet unclear.

**Figure 3.**
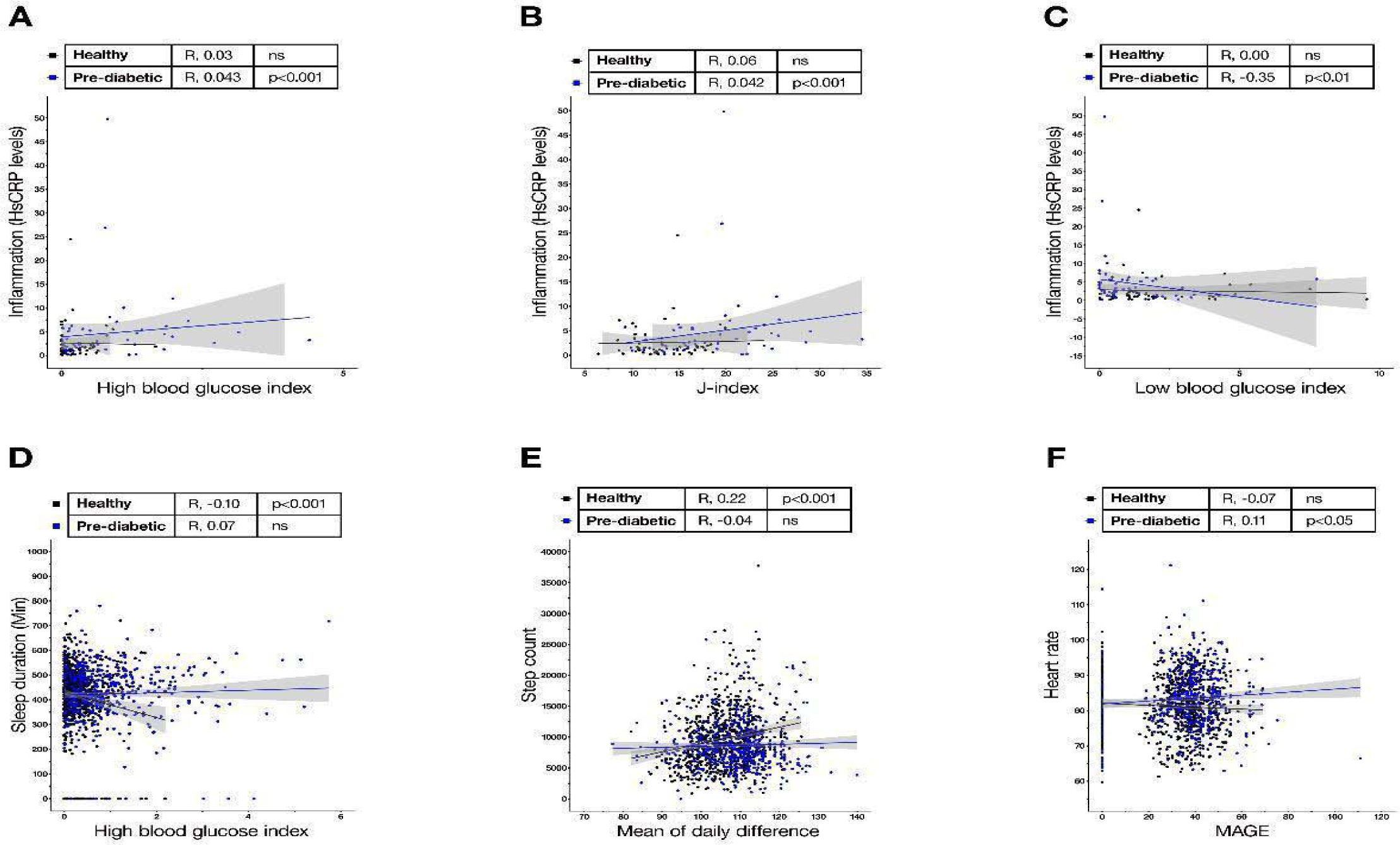
Secondary outcomes: Correlations of GV with clinical biomarkers associated with impaired glucose tolerance (IGT) and commonly tracked fitness measures. **R - correlation coefficient. Linked to Supplementary Tables 2-5**

Fitness tracker-derived motility and sleep metrics showed weak but significant correlations. There was a significant positive correlation between ADRR, MODD, and step count in non-diabetics (**Supplementary Table S5, Figure 3E**), while heart rate was positively correlated with ADRR and MAGE in pre-diabetics (**Supplementary Table 5, Figure 3F**).

### Correlations with CGM-derived GV indices with HOMA-IR, OGTT, and HbA1c

While CGM-derived indices have been utilized to develop algorithms to differentiate between healthy and diabetic individuals, studies evaluating the correlation of CGM-derived GV metrics with clinical gold standards to detect impaired glucose tolerance (IGT) and insulin resistance such as HbA1c, OGTT, and HOMA-IR are scarce.^31-33^ To bridge this gap, we carried out a post-hoc analysis, correlating the factors recorded in our study **(Table 1)**. For all clinical biomarkers, daily glycemic variability (J-index) was a strong associate in the pre-diabetes group with highly significant positive correlations. The HBGI count in both healthy and pre-diabetics was a consistent measure of elevated OGTT and HbA1c levels. However, HBGI correlated with increased HOMA IR showing that insulin resistance is a feature of IGT space and not a healthy glucose control space. LBGI showed some interesting correlations with OGTT and HOMA-IR in the healthy group, perhaps indicating that hypoglycemic events requiring glucose mobilization are more tightly regulated in normal glucose tolerance regimes. Specifically, in healthy participants, the glucose swing as captured by the ADRR or MODD is likely a better predictor of nascent insulin resistance. In summary, there is a difference in the type of glycemic parameters that a healthy or an IGT user of CGMs should focus on.

**Table 1.**
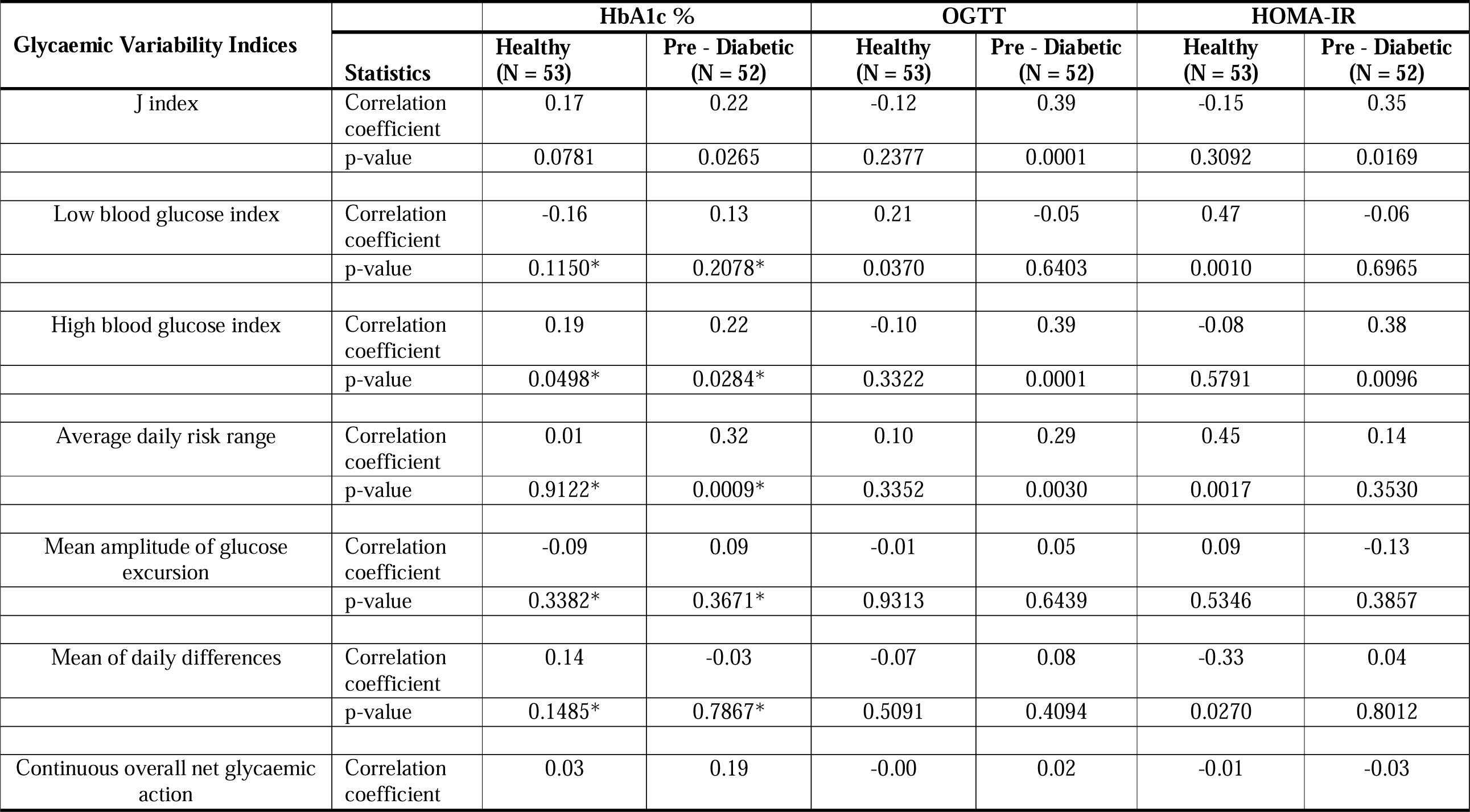
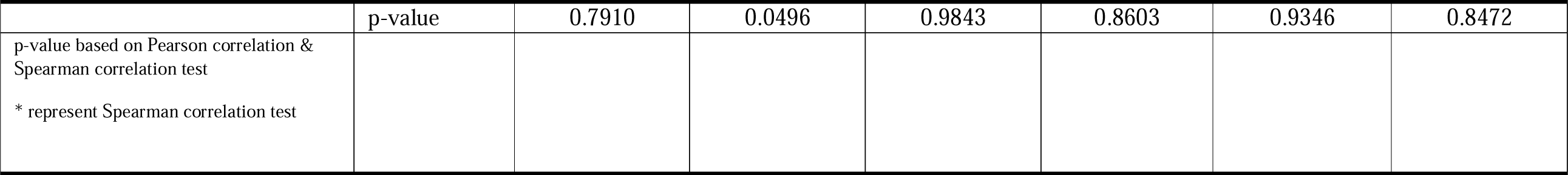
Correlation between Glycaemic variability indices and HbA1C, OGTT, and HOMA-IR (PP Population)

### Correlations of MS with biomarkers associated with metabolic syndrome and fitness metrics

The MS metric was developed as an all-encompassing snapshot of glucose tolerance and by extension, of the glycemic fitness of UH-M1 users. While this proprietary score is composed of weighted contributions from an individual’s glycemic variability, time in range, and mean glucose values, we tested the correlation of MS with the clinical biomarkers gathered for both pre- and non-diabetic groups, in an effort to benchmark in a controlled population. As shown in Table 2, MS had extremely strong negative correlations with inflammation (HsCRP), HbA1c, OGTT, and HOMA-IR in pre-diabetic participants. In non-diabetics, significant negative correlations were found between OGTT and HOMA-IR only. As a counterpoint, MS did not show any correlation with single-snapshot FBG levels in either group, attesting to its cumulative informational quality. In the fitness tracker metrics, the MS in both groups was weakly correlated with step count, heart rate, and sleep duration. Heart rate (being tightly regulated), displayed the expected negative correlation with MS and was statistically significant for both groups.

**Table 2:** Correlation Between Metabolic Score and Stress (by Cortisol), Inflammation (by Hs-CRP), HbA1C%, OGTT values, HOMA IR, fasting glucose levels, and fitness variables (PP Population)

| **Metabolic Score with**<br>**PSQI Score, Stress, Inflammation,**<br>**HbA1C %, OGTT, HOMA-IR and GMI** | **Statistics** | **Healthy**<br>**(N = 53)** | **Pre - Diabetic**<br>**(N = 52)** |
| --- | --- | --- | --- |
| Stress | Correlation coefficient | -0.05 | 0.16 |
|  | p-value | 0.6023* | 0.1142* |
| Inflammation | Correlation coefficient | -0.16 | -0.38 |
|  | p-value | 0.2748* | 0.0055* |
| HbA1C (%) | Correlation coefficient | -0.15 | -0.36 |
|  | p-value | 0.1397 | 0.0002 |
| OGTT | Correlation coefficient | -0.27 | -0.42 |
|  | p-value | 0.0056* | <0.0001* |
| HOMA-IR | Correlation coefficient | -0.37 | -0.43 |
|  | p-value | 0.0113* | 0.0036* |
| Fasting Blood Glucose | Correlation coefficient | 0.01 | -0.15 |
|  | p-value | 0.9186 | 0.1189 |
| Step Count | Correlation coefficient | -0.004 | 0.07 |
|  | p-value | 0.908 | 0.0978 |
| Heart Rate | Correlation coefficient | -0.13 | -0.08 |
|  | p-value | 0.0006 | 0.0380 |
| Sleep Duration | Correlation coefficient | 0.12 | -0.08 |
|  | p-value | 0.0014 | 0.0356 |
| p-value based on Pearson correlation & spearman correlation test. * represents Spearman Correlation test |  |  |  |

Interestingly, a small but significant positive correlation was found between sleep duration and MS in non-diabetics, with a paradoxical weak, negative correlation between sleep and MS in pre-diabetics.

## Discussion

The study cohort was representative of an urban, young adult Indian population who were non-obese, but overweight which constitutes a third of Indian adults as per the National Family Health Survey (NFHS-5) conducted in 2021.^34^ This is also the population most prone to developing prediabetes and diabetes in India, although the share of rural patients is seen to be increasing.^19-20^ To our knowledge, this is the first study that provides CGM-derived guidance values of glycemic indices and variability in non-diabetic (healthy) and pre-diabetic Indian populations. Furthermore, our extended analyses revealed that of the multiple glycemic indices used in this therapy area, there is a difference in the significance of correlating benchmarks of glucose control like Hba1c, OGTT, and insulin resistance by HOMA-IR between healthy and pre-diabetics. Interestingly, UH-MS mirrors many of the trends in glycemic dysfunction found in pre-diabetics and offers a dynamic, easy-to-understand metric that can generate personalized information. Finally, we report that inflammation had the strongest positive correlation with glycemic indices in pre-diabetics indicating that significant metabolic dysregulation is already underway in this group. This supports the notion that glucose tolerance regimes are most likely a contiguous spectrum, rather than discrete states of non-diabetes, pre-diabetes, and diabetes.

In terms of digital health tracking, studies like GLITTER, Twin Precision Nutrition (TPN) Program, and ambulatory glucose profiling (AGP) have demonstrated the power of CGM-based tracking in making real-time interventional decisions like dietary- or exercise changes, dosage changes of insulin, etc, that can be tracked by patients and clinicians simultaneously.^35-37^ Within India, electronic health (e-health) and mobile health (m-health) initiatives have been successfully used to provide support, motivation, and directional suggestions to large cohorts to make healthier lifestyle choices.^38-39^ Internationally, large cohort studies have been undertaken in developed countries like the Dehgani-Zahedani et al, 2021 (Sugar.AI initiative), that show that a 10-day CGM app-based tracking regime can significantly promote healthier metabolic-oriented choices in healthy and at-risk individuals.^40^ The gap lay in reference data of CGM-derived glycemic metrics for non-diabetic/healthy Indians (and by extension South Asians), which were either not the focus of clinical studies like GLITTER and Twin, or underrepresented in North American and European studies. With a demographic contribution of over a fifth of the world’s population, and being a high-risk group for developing metabolic syndromes, this group represents an important resource for gathering natural history, baseline evidence development, and increased surveillance.

In terms of primary outcomes of the study, we found a consistent and significant difference in mean glucose levels between non-diabetics and pre-diabetics with no significant differences in TIR per the broader ADA guideline range. Interestingly, the rTIR per app guidance of a target range of 70-110mg/dL was significantly improved in both groups and across all days, highlighting the need to adjust metrics to derive useable information in a context-specific manner. The improved rTIR was also tracked with lesser hyper- and hypo-glycemic events in both groups over days. CGM-based metrics of GV as measured by SD, CV, and MAGE also showed significant improvement over time in both groups. However, the data alludes to the fact that for more lasting changes in FBG and allied indices, tracking for a period longer than 14 days might be required to yield lasting corrective changes for pre-diabetics.

Inflammation as measured by hsCRP had the strongest positive correlation with glycemic indices in pre-diabetics. This confirms the fact that there is already significant metabolic dysregulation in this group. This is of particular importance as a clear association between cardiovascular disease and prediabetes has emerged over the past few years.^41^ Furthermore, Indians have been known to have a higher hsCRP level both in healthy and pre-diabetics.^42-43^ In this study, pre-diabetics displayed a trend of having roughly twice the levels of inflammation as compared to healthy participants even though the cohort was comparably overweight. This is in dramatic contrast to the cortisol data, which seems to indicate that stress levels were comparable between the groups.

In the domains of sleep duration, heart rate, and step count, the data indicated that glycemic indices were only correlated in the healthy group. The sleep duration correlates reported in this study were derived using the FitBit tracker. Results indicate that higher GV and increased episodes of hyperglycemia have noticeably less impact on sleep duration in healthy people and are more of a concern for at-risk individuals such as pre-diabetics. Hence these metrics take on added significance only when a pre-diabetes diagnosis has been made. Although not in the remit of this current study, it is possible that multi-modal analyses of data for pre-diabetics could identify sub-populations with specific sleep disturbance patterns that correlate better with impaired glucose control.

Step count and heart rate monitoring have now become integral parts of any fitness agenda. Although the strength of the correlations between these factors and GV was found to be weak in this study, it should be considered that other coexisting factors may also influence the relationship. For healthy individuals, a recent study reported every 1000 step increase per day was correlated with a blunted GV the following day but not within the same day.^44^ Per protocol, this study analysed within-day GV with step count and heart rate which may be the reason for the weaker association of these metrics in our study. Nevertheless, the positive correlation of step count with mean daily differences is a useful indicator of daily swings which can potentially be leveraged to optimally fuel for exercise sessions in healthy individuals.

An important post-hoc analysis conducted on the study data aimed to address a vital gap in point-of-care surveillance. With busy lifestyles, people often miss conducting wellness check-ups involving gold-standard biomarkers (HbA1c, OGTT, and HOMA-IR) for glucose tolerance. Hence, by the time systemic symptoms appear, an individual has already progressed to an advanced clinical state within the diabetic spectrum. Recent studies have investigated the predictive power of CGM systems to differentiate healthy, pre-diabetic, and diabetic individuals using these biomarkers.^33,45^ Our results indicate that daily GV (J-index) was a strong proxy in the pre-diabetes group of all three clinical parameters in the Indian and South Asian demographics. This relationship did not hold true in the healthy participant group indicating a need to attribute differential weightage to these indices based on diagnosis. The HBGI in both healthy and pre-diabetics was a consistent measure of elevated OGTT and HbA1c. However, hyperglycaemic events correlated with increased HOMA IR indicating that insulin resistance was a feature of the IGT space specifically. Instead, in healthy participants, the glucose swing daily as captured by the ADRR or MODD was a better predictor of insulin resistance. We hope that these relationships will be explored in larger controlled and real-world cohorts to determine the best continuous proxies for clinical biomarkers of glucose dysfunction.

Wellness/risk score calculators offer an easy-to-understand metric for laypeople to appreciate the risk of developing various conditions. Scores such as the Life’s Essential 8 ^TM^ developed by the American Heart Association predict cardiovascular health based on self-reported responses to generalized questions and offer a demographic risk factor-powered perspective to an individual’s heart health. ^46^ On the other hand, personalized wearables and digital health monitoring devices are accompanied by aggregate, algorithmic scores to serve as an easy-to-understand handle for a user to track his/her health and potentially modify their behaviour. Only a handful of these scores have been validated using cohorts in controlled settings and benchmarked to accepted clinical biomarkers. To our knowledge, our study is novel in its approach to clinically validate UH-MS and the results indicate that the score is an effective digital proxy for IGT and insulin resistance in the population studied. Given its relevance to glucose tolerance and insulin resistance, it is plausible to imagine an expanded use case in other ethnic groups.

The main limitation of the study was the short duration of CGM use (14-day period) which may have been minimally sufficient to glean early indications of the positive impact of UH-M1-based tracking. A longer observational study is required to derive more substantive conclusions on app-guided lifestyle changes or the data mining of real-world, non-controlled evidence from the UH user community. Other limitations included capturing basic sleep duration using the Fitbit activity tracker, which did not have refined measurement of epochs, and REM vs. non-REM which could have linked more accurately to GV measured in pre-diabetics. This also holds true for opposing correlation patterns of sleep duration with metabolic score. It is not clear how a high MS (indicative of strong glycemic control) is linked with poor sleep in pre-diabetics at the same time that good sleep is coincident with high MS in non-diabetics.

With this study, we lay the foundation for further exploring valuable dynamics of GV with day-to-day activities as well as the metabolic parameters in non-diabetic and pre-diabetic diabetic individuals. The findings underscore the value of using CGMs for wellness and preventive surveillance. Long-term studies will provide more data on these associations and may serve as a guide to managing these individuals by making adequate lifestyle and if required, pharmacological changes.

## Conclusions

This study provides a reference data set for CGM-derived glycemic metrics in adult Indian/South Asian groups, for a 14-day use period. The data indicates that the specific GV indices correlate better with clinical biomarkers and lifestyle indicators for pre-diabetics as compared to non-diabetics and should be given varying weightage by clinicians when reviewing the fitness of the two groups. Finally, the study also provides critical support for the use of MS score as a digital proxy for glycemic health.

## Supporting information

STROBE checklist

Trial registry snapshot

## Declarations

### Ethical approval

Ethical approval was obtained from each of the clinical trials individually. This is captured in Supplementary Protocol file uploaded and information is available publicly on the Clinical Trial Registry of India website, accessed by trial identifier CTRI/2022/08/044808

### Guarantor

B.S. (ORCID ID: 0009-0009-4865-380X)

### Author Contributions

B.S.: Conceptualization, Methodology, Overall supervision and coordination, and manuscript review; M.C.-Trial coordination, Curation of Data, Extended data analysis, Visualization, manuscript review; M.K. and V.S.-Conceptualization, Funding acquisition, Technological development, and support.

All named authors meet the International Committee of Medical Journal Editors (ICMJE) criteria for authorship for this article, take responsibility for the integrity of the work, and have given their approval for this version to be published.

## Acknowledgments

Acknowledgments: The authors would like to thank all the participants in the study. Authors are grateful to Dr. Prabhat Ranjan Sinha, Dr. R.P. Rajesh, Dr. Neeta Deshpande, Dr. Suresh S.M., Dr. Banshi Saboo, Dr. Hamsraj Alva, Dr. B.V.S.N. Raju, Dr. Sriharee Kulkarni, and Dr. Pankaj Aneja who were principal investigators at the various trial sites. Additionally, the authors acknowledge the contributions of Triomics Healthcare for trial automation and trial management, Dr. Aditi Bhattacharya for scientific analyses and publication development support, and Dr. Aafrin Khan for editorial support of the manuscript.

## Data availability

Study outcomes data can be made available upon reasonable request. The M1 and MS platform codes and technical details are proprietary assets of Ultrahuman and will not be disclosed.

## Supplementary section

**Supplementary Figure S1:**
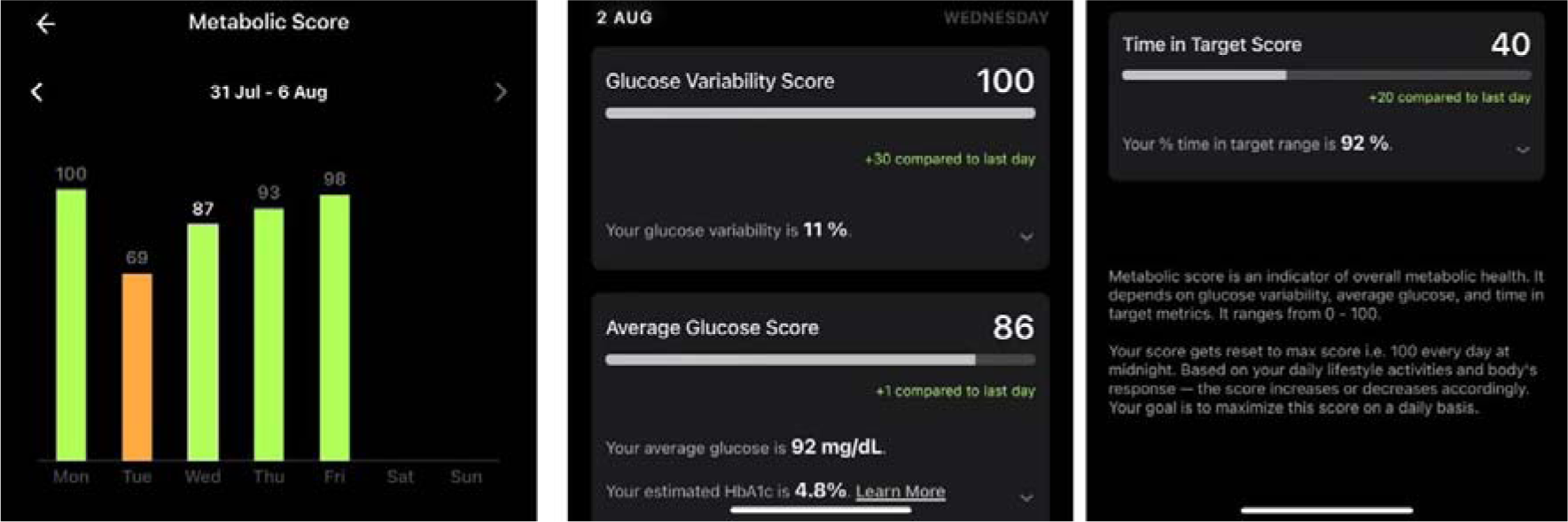
Exemplar snapshot of the MS information panel on the M1 platform.

**Supplementary Figure S2:**
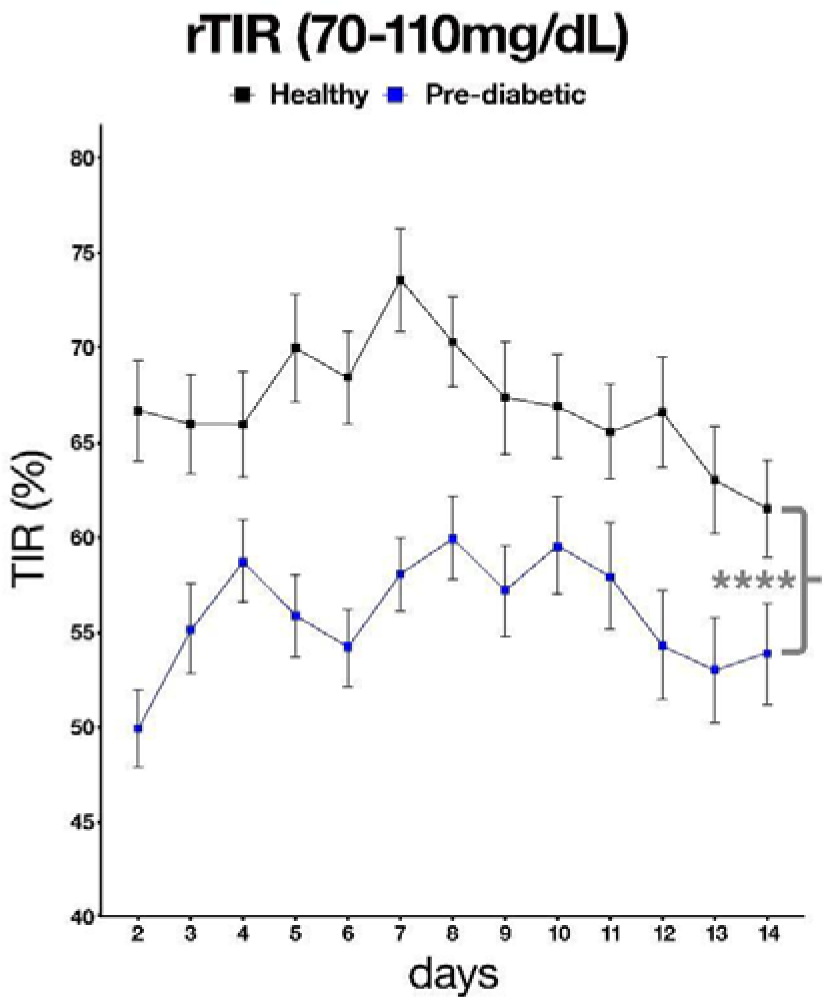
Restricted time in range in healthy vs pre-diabetic within the stipulated time-frame. **rTIR: restricted Time in range, Statistical analyses: Two-factor ANOVA** (cohort x day; main effect, cohort: p<0.0001; main effect, day: p<0.0001, interaction cohort x day: p<0. 0.00001)

**Supplementary Table S1:** Baseline demographics.

| Parameter | Statistics | Healthy<br>(N=53) | Pre-Diabetic<br>(N=52) |
| --- | --- | --- | --- |
| Gender n (%) | Male | 39 (73.6) | 45 (86.5) |
| Age (Years) | N* | 53 | 52 |
|  | Mean (SD) | 32.8 (6.56) | 35.6 (6.94) |
|  | Median (Q1, Q3) | 30.0 (28.0, 37.0) | 35.5 (30.5, 41.0) |
|  | Min, Max | 25.0, 47.0 | 25.0, 49.0 |
| BMI (kg/m<sup>2</sup>) | N* | 53 | 52 |
|  | Mean (SD) | 25.571 (2.8634) | 26.456 (2.6311) |
|  | Median (Q1, Q3) | 25.390 (24.050, 27.940) | 26.830 (24.905, 28.410) |
|  | Min, Max | 20.030, 29.760 | 20.260, 29.940 |

BMI: Body mass index; SD: Standard deviation.

**Supplementary Table S2:** Correlation between Glycaemic variability indices and inflammation (as measured by Hs-CRP) (PP ulation)

| Inflammation with<br>glycaemic variability indices | Statistics | Healthy<br>(N = 53) | Pre - Diabetic<br>(N = 52) |
| --- | --- | --- | --- |
| J index | Correlation coefficient | 0.06 | 0.42 |
|  | p-value | 0.6959 | 0.0020 |
| Low blood glucose index | Correlation coefficient | -0.002 | -0.35 |
|  | p-value | 0.9876 | 0.0108 |
| High blood glucose index | Correlation coefficient | 0.03 | 0.43 |
|  | p-value | 0.8162 | 0.0016 |
| Average daily risk range | Correlation coefficient | -0.05 | -0.12 |
|  | p-value | 0.7448 | 0.3840 |
| Mean amplitude of glucose excursion | Correlation coefficient | 0.01 | -0.10 |
|  | p-value | 0.9637 | 0.4692 |
| Mean of daily differences | Correlation coefficient | -0.10 | 0.20 |
|  | p-value | 0.5034 | 0.1492 |
| Continuous overall net glycaemic action | Correlation coefficient | 0.04 | 0.004 |
|  | p-value | 0.7912 | 0.9772 |
| p-value based on Pearson correlation & spearman correlation test<br>* represent spearman correlation test |  |  |  |

**Supplementary Table S3:** Correlation between glycaemic variability indices and stress (as measured by cortisol) (PP Population)

| **Stress with**<br>**glycaemic variability indices** | **Statistics** | **Healthy**<br>**(N = 53)** | **Pre - Diabetic**<br>**(N = 52)** |
| --- | --- | --- | --- |
| J index | Correlation coefficient | 0.06 | -0.15 |
|  | p-value | 0.5615* | 0.1253* |
| Low blood glucose index | Correlation coefficient | -0.04 | 0.09 |
|  | p-value | 0.7242* | 0.3663* |
| High blood glucose index | Correlation coefficient | 0.04 | -0.16 |
|  | p-value | 0.6688* | 0.0970* |
| Average daily risk range | Correlation coefficient | 0.01 | -0.08 |
|  | p-value | 0.8956* | 0.4105* |
| Mean amplitude of glucose excursion | Correlation coefficient | -0.09 | 0.06 |
|  | p-value | 0.3760* | 0.5502* |
| Mean of daily differences | Correlation coefficient | -0.07 | -0.08 |
|  | p-value | 0.4685* | 0.4078* |
| Continuous overall net glycaemic action | Correlation coefficient | -0.09 | -0.05 |
|  | p-value | 0.3521 | 0.5844 |
| p-value based on Pearson correlation & spearman correlation test<br>* represent spearman correlation test |  |  |  |

**Supplementary Table S4:** Correlation between Glycaemic variability indices and sleep duration (PP Population)

| **Glycaemic variability indices** | **Statistics** | **Healthy**<br>**(N = 53)** | **Pre - Diabetic**<br>**(N = 52)** |
| --- | --- | --- | --- |
| J index | Correlation coefficient | -0.08 | 0.07 |
|  | p-value | 0.0336 | 0.0872 |
| Low blood glucose index | Correlation coefficient | 0.01 | -0.05 |
|  | p-value | 0.8938 | 0.2408 |
| High blood glucose index | Correlation coefficient | -0.10 | 0.07 |
|  | p-value | 0.0082 | 0.0845 |
| Average daily risk range | Correlation coefficient | -0.07 | 0.06 |
|  | p-value | 0.0768 | 0.1262 |
| Mean amplitude of glucose excursion | Correlation coefficient | -0.06 | -0.06 |
|  | p-value | 0.1025 | 0.1445 |
| Mean of daily differences | Correlation coefficient | -0.05 | -0.04 |
|  | p-value | 0.1591 | 0.3318 |
| Continuous overall net glycaemic action | Correlation coefficient | -0.09 | -0.10 |
|  | p-value | 0.0159 | 0.0115 |
| p-value based on Pearson correlation & spearman correlation test<br>* represent spearman correlation test |  |  |  |

**Supplementary Table S5:**
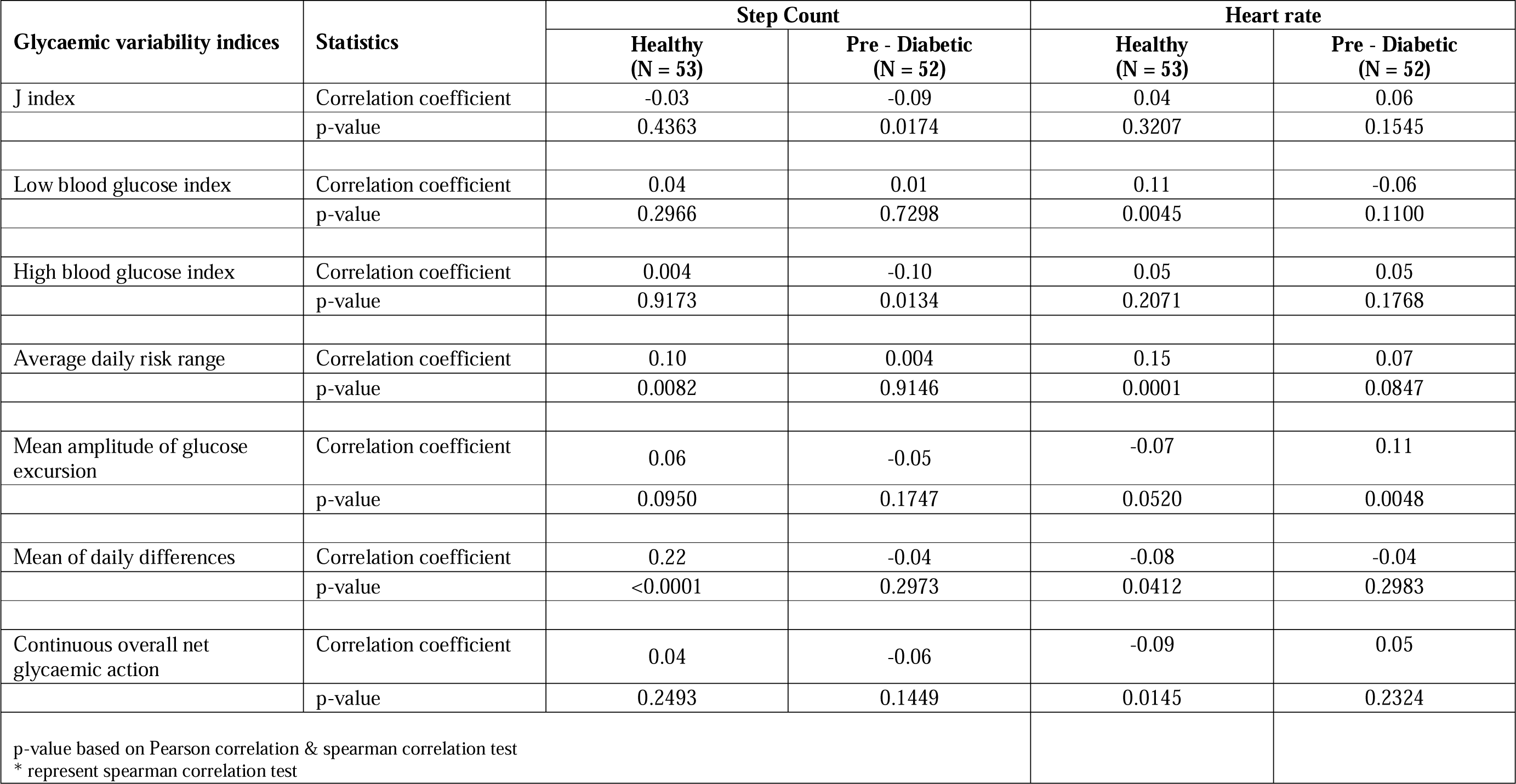
Correlation between Glycaemic variability indices and physical activity (step count) (PP Population)

**Supplementary Table 7:** Details of trial sites, principal investigators, ethics committee approval dates.

| **Site Name (Name of Ethics Board)** | **Site Address** | **Date of Approval** |
| --- | --- | --- |
| Aakash Healthcare Private Limited (Hospital Ethics Board) | Aakash Healthcare, Private Limited Hospital Plot Road No. 201, Sector-3, Dwarka, New Delhi -110075 | 11<sup>th</sup> August 2022 |
| Aadhavvan Diabetes & Research Centre (Universal Ethics Committee) | No.3, 5th Street, Eswar Nagar, Kodambakkam, Chennai, Tamil Nadu - 600024 | 08<sup>th</sup> August 2022 |
| Belgaum Diabetes Centre (Diabetes Centre Ethics Committee) | Ground and second floor, Maruti street, Belgaum, Karnataka- 590001 | 12<sup>th</sup> August 2022 |
| Suresh Diacare (Bangalore Ethics Committee) | No 723B, 11th Main Rd, 3rd Block, Rajajinagar, Bengaluru, Karnataka 560010 | 26<sup>th</sup> September 2022 |
| Diacare Research (Shrey Hospital Institutional Ethics Committee) | 1,2 Gandhi Park, near Nehrunagar, Ambawadi, Ahmedabad, Gujarat- 380015 | 12<sup>th</sup> August 2022 |
| Vinaya Hospital, (Hospital Ethics Committee) | Vinaya Hospital and Research Centre (a unit of KIMS), Karangalpady, Mangaluru, Karnataka-575003 | 30<sup>th</sup> July 2022 |
| Induss Hospital, (Institutional Ethics Committee) | Induss Hospital, Opp. Kothapet Fruit Market Kothapet, Sri Sai Shivani Complex, HUDA Complex, Saroornagar, Hyderabad, Telangana- 500035 | 14<sup>th</sup> October 2022 |
| Kulkarni's Medzonne (Bangalore Ethics Committee) | Kulkarni's Medzonne, GD Naidu Hall, Mohan Matrix, 450<sup>th</sup>, 12<sup>th</sup> cross road, Near Vidya Bharti School, Mahalakshmiapuram, Bengaluru, Karnataka. 560086 | 03<sup>rd</sup> October 2022 |
| Naveda Healthcare Centre (Good Society Ethical Research Oversight Committee) | A-1/81, Sector-8, Rohini, Delhi. 110085 | 02<sup>nd</sup> August 2022 |

## References

1. World health organization-https://www.who.int/health-topics/diabetes#tab=tab_1 (Accessed 23rd July 2023)

2. Yip WCY, Sequeira IR, Plank LD, and Poppitt SD. Prevalence of Pre-Diabetes across Ethnicities: A Review of Impaired Fasting Glucose (IFG) and Impaired Glucose Tolerance (IGT) for Classification of Dysglycaemia. Nutrients. 2017; 22;9(11):1273.

3. Saeedi P, Petersohn I, Salpea P, et al. IDF Diabetes Atlas Committee. Global and regional diabetes prevalence estimates for 2019 and projections for 2030 and 2045: Results from the International Diabetes Federation Diabetes Atlas, 9th edition. Diabetes Res Clin Pract. 2019;157:107843.

4. Dali-Youcef N, Mecili M, Ricci R, and Andrès E. Metabolic inflammation: connecting obesity and insulin resistance. Ann Med. 2013; 45(3):242–53.

5. Fazli GS, Moineddin R, Bierman AS, and Booth GL. Ethnic variation in the conversion of prediabetes to diabetes among immigrant populations relative to Canadian-born residents: a population-based cohort study. BMJ Open Diabetes Res Care. 2020; 8(1):e000907.

6. Tuomilehto J, Lindström J, Eriksson JG, et al. Finnish Diabetes Prevention Study Group. Prevention of type 2 diabetes mellitus by changes in lifestyle among subjects with impaired glucose tolerance. N Engl J Med. 2001; 344(18):1343-50.

7. Prevention or Delay of Type 2 Diabetes: Standards of Medical Care in Diabetes—2021. American Diabetes Association. Diabetes J. Diabetes Care 2021;44(Supplement_1):S34–S39

8. Duan D, Kengne AP, and Echouffo-Tcheugui JB. Screening for Diabetes and Prediabetes. Endocrinol Metab Clin North Am. 2021; 50(3):369–385.

9. Bergman M, Abdul-Ghani M, DeFronzo RA, et al. Review of methods for detecting glycemic disorders. Diabetes Res Clin Pract. 2020; 165:108233.

10. Rodbard D. Continuous Glucose Monitoring: A Review of Successes, Challenges, and Opportunities. Diabetes Technol Ther. 2016; 18 Suppl 2(Suppl 2):S3-S13.

11. Soliman A, DeSanctis V, Yassin M, et al. Continuous glucose monitoring system and new era of early diagnosis of diabetes in high risk groups. Indian J Endocrinol Metab. 2014; 18(3):274–82.

12. Klonoff DC, Nguyen KT, Xu NY, et al. Use of Continuous Glucose Monitors by People Without Diabetes: An Idea Whose Time Has Come? J Diabetes Sci Technol. 2022; doi: 10.1177/19322968221110830.

13. Daly A, and Hovorka R. Technology in the management of type 2 diabetes: Present status and future prospects. Diabetes Obes Metab. 2021; 23(8):1722–1732.

15. https://blog.ultrahuman.com/blog/beginners-guide-to-the-ultrahuman-m1/ (accessed 23rd July 2023)

16. https://blog.ultrahuman.com/blog/how-is-your-metabolic-score-calculated/ (Accessed 13th August 2023)

17. Mohan V, Joshi S, Mithal A, et al. Expert Consensus Recommendations on Time in Range for Monitoring Glucose Levels in People with Diabetes: An Indian Perspective. Diabetes Ther. 2023; 14(2):237–249.

18. Wells JC, Pomeroy E, Walimbe SR, et al. The Elevated Susceptibility to Diabetes in India: An Evolutionary Perspective. Front Public Health. 2016; 4:145.

19. Anjana RM, Unnikrishnan R, Deepa M, et al. ICMR-INDIAB Collaborative Study Group. Metabolic non-communicable disease health report of India: the ICMR-INDIAB national cross-sectional study (ICMR-INDIAB-17). Lancet Diabetes Endocrinol. 2023; 11(7):474-489.

20. Anjana RM, Deepa M, Pradeepa R et al. ICMR–INDIAB Collaborative Study Group. Prevalence of diabetes and prediabetes in 15 states of India: results from the ICMR-INDIAB population-based cross-sectional study. Lancet Diabetes Endocrinol. 2017; 5(8):585-596.

21. Anjana RM, Shanthi Rani CS, et al. Incidence of Diabetes and Prediabetes and Predictors of Progression Among Asian Indians: 10-Year Follow-up of the Chennai Urban Rural Epidemiology Study (CURES). Diabetes Care. 2015; 38(8):1441–8.

22. https://www.accessdata.fda.gov/cdrh_docs/pdf19/K193371.pdf (Accessed 21st July 2023)

23. Buysse DJ, Reynolds CF 3rd, Monk TH, et al. The Pittsburgh Sleep Quality Index: a new instrument for psychiatric practice and research. Psychiatry Res. 1989; 28(2):193–213.

24. Bailey T, Bode BW, Christiansen MP, et al. The Performance and Usability of a Factory-Calibrated Flash Glucose Monitoring System. Diabetes Technol Ther. 2015; 17(11):787–94.

25. https://www.R-project.org. R Core Team (2021). R: A language and environment for statistical computing. R Foundation for Statistical Computing, Vienna, Austria. (Accessed March 2023).

26. Battelino T, Danne T, Bergenstal RM, et al. Clinical Targets for Continuous Glucose Monitoring Data Interpretation: Recommendations from the International Consensus on Time in Range. Diabetes Care. 2019; 42(8):1593–1603.

27. Hill NR, Oliver NS, Choudhary P, et al. Normal reference range for mean tissue glucose and glycemic variability derived from continuous glucose monitoring for subjects without diabetes in different ethnic groups. Diabetes Technol Ther. 2011; 13(9):921–8.

28. Jaiswal A, Tabassum R, Podder A, et al. Elevated level of C-reactive protein is associated with risk of prediabetes in Indians. Atherosclerosis. 2012; 222(2):495–501.

29. Sonnier T, Rood J, Gimble JM, and Peterson CM. Glycemic control is impaired in the evening in prediabetes through multiple diurnal rhythms. J Diabetes Complications. 2014; 28(6):836–43.

30. Hur MH, Lee MK, Seong K, and Hong JH. Deterioration of Sleep Quality According to Glycemic Status. Diabetes Metab J. 2020; 44(5):679–686.

31. Mishra A, Podder V, Modgil S, et al. Higher Perceived Stress and Poor Glycemic Changes in Prediabetics and Diabetics Among Indian Population. J Med Life. 2020;13(2):132–137.

32. Satya Krishna SV, Kota SK, Modi KD. Glycemic variability: Clinical implications. Indian J Endocrinol Metab. 2013;17(4):611–9.

33. Acciaroli G, Sparacino G, Hakaste L, et al. Diabetes and Prediabetes Classification Using Glycemic Variability Indices From Continuous Glucose Monitoring Data. J Diabetes Sci Technol. 2018;12(1):105–113.

34. main.mohfw.gov.in/sites/default/files/NFHS-5_Phase-II_0.pdf (Accessed 22nd July 2023)

35. Jain AB. Glycemic improvement with a novel interim intervention technique using retrospective professional continuous glucose monitoring (GLITTER study): A study from Mumbai, India. Diabetes Metab Syndr. 202;15(3):703–709.

36. Shamanna P, Saboo B, Damodharan S, et al. Reducing HbA1c in Type 2 Diabetes Using Digital Twin Technology-Enabled Precision Nutrition: A Retrospective Analysis. Diabetes Ther. 2020; 11(11):2703–2714.

37. Anjana RM, Kesavadev J, Neeta D, et al. Multicenter Real-Life Study on the Effect of Flash Glucose Monitoring on Glycemic Control in Patients with Type 1 and Type 2 Diabetes. Diabetes Technol Ther. 2017; 19(9):533–540.

38. Ranjani H, Nitika S, Anjana R, et al. Impact of noncommunicable disease text messages delivered via an app in preventing and managing lifestyle diseases: results of the myArogya worksite-based effectiveness study from India. J Diabetol. 2020; 11: 90–100.

39. Pfammatter A, Spring B, Saligram N, et al. mHealth Intervention to Improve Diabetes Risk Behaviors in India: A Prospective, Parallel Group Cohort Study. J Med Internet Res. 2016; 18(8):e207. doi: 10.2196/jmir.5712.

40. Dehghani Zahedani A, Shariat Torbaghan S, Rahili S, et al. Improvement in Glucose Regulation Using a Digital Tracker and Continuous Glucose Monitoring in Healthy Adults and Those with Type 2 Diabetes. Diabetes Ther. 2021;12(7):1871–1886.

41. Zand A, Ibrahim K, Patham B. Prediabetes: Why Should We Care? Methodist Debakey Cardiovasc J. 2018; 14(4):289–297.

42. Kamath DY, Xavier D, Sigamani A, and Pais P. High sensitivity C-reactive protein (hsCRP) & cardiovascular disease: An Indian perspective. Indian J Med Res. 2015; 142(3):261–8.

43. Ghule A, Kamble TK, Talwar D, et al. Association of Serum High Sensitivity C-Reactive Protein With Pre-diabetes in Rural Population: A Two-Year Cross-Sectional Study. Cureus. 2021;13(10):e19088. doi: 10.7759/cureus.19088.

44. El Fatouhi D, Héritier H, Allémann C, et al. Associations Between Device-Measured Physical Activity and Glycemic Control and Variability Indices Under Free-Living Conditions. Diabetes Technol Ther. 2022; 24(3):167–177.

45. Longato E, Acciaroli G, Facchinetti A, et al. Simple Linear Support Vector Machine Classifier Can Distinguish Impaired Glucose Tolerance Versus Type 2 Diabetes Using a Reduced Set of CGM-Based Glycemic Variability Indices. J Diabetes Sci Technol. 2020; 14(2):297–302.

46. https://www.heart.org/en/healthy-living/healthy-lifestyle/lifes-essential-8. (Accessed 13th August 2023)

