## Supplementary material for "Metabolic health tracking using Ultrahuman M1 continuous glucose monitoring platform in non- and pre-diabetic Indians: a multi-armed observational study": STROBE checklist

| | Item No. | Recommendation | Page No. | Relevant text from manuscript |
|---|---|---|---|---|
| **Title and abstract** | 1 | (*a*) Indicate the study's design with a commonly used term in the title or the abstract | 1 | Metabolic health tracking using Ultrahuman M1 continuous glucose monitoring platform in non- and pre-diabetic Indians: a multi-armed observational study |
| | | (*b*) Provide in the abstract an informative and balanced summary of what was done and what was found | 1 | Abstract: lines 17-42. |
| **Introduction** | | | | |
| Background/rationale | 2 | Explain the scientific background and rationale for the investigation being reported | 4-5 | Line 58-65, 69-79 |
| Objectives | 3 | State specific objectives, including any prespecified hypotheses | 5 | Line 107-112 |
| **Methods** | | | | |
| Study design | 4 | Present key elements of study design early in the paper | 6 | Line 114-143 |
| Setting | 5 | Describe the setting, locations, and relevant dates, including periods of recruitment, exposure, follow-up, and data collection | 6-7, | Line 114-143, 146-178, Supplementary Table 7 |
| Participants | 6 | (*a*) *Cohort study*—Give the eligibility criteria, and the sources and methods of selection of participants. Describe methods of follow-up<br>*Case-control study*—Give the eligibility criteria, and the sources and methods of case ascertainment and control selection. Give the rationale for the choice of cases and controls<br>*Cross-sectional study*—Give the eligibility criteria, and the sources and methods of selection of participants | 7 & 8 | Line 146-178 |
| | | (*b*) *Cohort study*—For matched studies, give matching criteria and number of exposed and unexposed<br>*Case-control study*—For matched studies, give matching criteria and the number of controls per case | NA | NA |

| | | | | |
|---|---|---|---|---|
| Variables | 7 | Clearly define all outcomes, exposures, predictors, potential confounders, and effect modifiers. Give diagnostic criteria, if applicable | 9 | Line 181-198 |
| Data sources/ measurement | 8* | For each variable of interest, give sources of data and details of methods of assessment (measurement). Describe comparability of assessment methods if there is more than one group | 9-10 | Line 201-221 |
| Bias | 9 | Describe any efforts to address potential sources of bias | NR | Observational study |
| Study size | 10 | Explain how the study size was arrived at | Supplementary file: Protocol | Section 6.1 |

Continued on next page

| | | | | |
|---|---|---|---|---|
| Quantitative variables | 11 | Explain how quantitative variables were handled in the analyses. If applicable, describe which groupings were chosen and why | 9-10 | Line 201-221 |
| Statistical methods | 12 | (*a*) Describe all statistical methods, including those used to control for confounding | 9-10 | Line 201-221 |
| | | (*b*) Describe any methods used to examine subgroups and interactions | 9-10 | Line 201-221 |
| | | (*c*) Explain how missing data were addressed | NA, 11 | Line 225-227 |
| | | (*d*) *Cohort study*—If applicable, explain how loss to follow-up was addressed<br>*Case-control study*—If applicable, explain how matching of cases and controls was addressed<br>*Cross-sectional study*—If applicable, describe analytical methods taking account of sampling strategy | NR | NR |
| | | (*e*) Describe any sensitivity analyses | NA | NA |
| **Results** | | | | |
| Participants | 13* | (a) Report numbers of individuals at each stage of study—eg numbers potentially eligible, examined for eligibility, confirmed eligible, included in the study, completing follow-up, and analysed | 11 | Line 225-227, figure 1 |
| | | (b) Give reasons for non-participation at each stage | 8 | Line 176-178 |
| | | (c) Consider use of a flow diagram | - | Figure 1 |
| Descriptive data | 14* | (a) Give characteristics of study participants (eg demographic, clinical, social) and information on exposures and potential confounders | 11 | Line 225-234 and Supplementary table S1 |
| | | (b) Indicate number of participants with missing data for each variable of interest | NA | NA |
| | | (c) *Cohort study*—Summarise follow-up time (eg, average and total amount) | NA | NA |
| Outcome data | 15* | *Cohort study*—Report numbers of outcome events or summary measures over time | 11-16 | Line 224-357 |
| | | *Case-control study*—Report numbers in each exposure category, or summary measures of exposure | | |
| | | *Cross-sectional study*—Report numbers of outcome events or summary measures | | |
| Main results | 16 | (*a*) Give unadjusted estimates and, if applicable, confounder-adjusted estimates and their precision (eg, 95% confidence interval). Make clear which confounders were adjusted for and why they were included | NA | NA |
| | | (*b*) Report category boundaries when continuous variables were categorized | 11-16 | Line 224-357 |
| | | (*c*) If relevant, consider translating estimates of relative risk into absolute risk for a meaningful time period | NA | NA |

Continued on next page

| | | | | |
|---|---|---|---|---|
| Other analyses | 17 | Report other analyses done—eg analyses of subgroups and interactions, and sensitivity analyses | 12, 15-16 | Lines: 253-258, 324-357 |
| **Discussion** | | | | |
| Key results | 18 | Summarise key results with reference to study objectives | 16 | Line 364-375 |
| Limitations | 19 | Discuss limitations of the study, taking into account sources of potential bias or imprecision. Discuss both direction and magnitude of any potential bias | 20-21 | Line: 461-471 |
| Interpretation | 20 | Give a cautious overall interpretation of results considering objectives, limitations, multiplicity of analyses, results from similar studies, and other relevant evidence | 21 | Line: 472-483 |
| Generalisability | 21 | Discuss the generalisability (external validity) of the study results | 21 | Line: 472-483 |
| **Other information** | | | | |
| Funding | 22 | Give the source of funding and the role of the funders for the present study and, if applicable, for the original study on which the present article is based | 21 | Line: 489 |

*Give information separately for cases and controls in case-control studies and, if applicable, for exposed and unexposed groups in cohort and cross-sectional studies.
