## Supplementary material for "Metabolic health tracking using Ultrahuman M1 continuous glucose monitoring platform in non- and pre-diabetic Indians: a multi-armed observational study": Trial registry snapshot

### Clinical Trial Details (PDF Generation Date :- Sat, 16 Sep 2023 00:13:04 GMT)

| | |
|---|---|
| **CTRI Number** | CTRI/2022/08/044808 [Registered on: 22/08/2022] - **Trial Registered Prospectively** |
| **Last Modified On** | 23/03/2023 |
| **Post Graduate Thesis** | No |
| **Type of Trial** | Observational |
| **Type of Study** | Cohort Study |
| **Study Design** | Other |
| **Public Title of Study** | Development of an Algorithm for Ultrahuman Health Platform |
| **Scientific Title of Study** | Development of an Algorithm to Assess Metabolic Health of Non-diabetic and Pre-diabetic South-Asian Population Using Continuous Glucose Monitoring Systems and Ultrahuman Health Platform |

**Secondary IDs if Any**

| Secondary ID | Identifier |
|---|---|
| NIL | NIL |

**Details of Principal Investigator or overall Trial Coordinator (multi-center study)**

| Details of Principal Investigator | |
|---|---|
| **Name** | Suresh Kumar Karri |
| **Designation** | Sr. Project Manager |
| **Affiliation** | Triomics Healthcare Pvt Ltd |
| **Address** | Level 5, Green Boulevard, Block C, Sector 62 Noida Uttar Pradesh 201 301 India<br>Gautam Buddha Nagar<br>UTTAR PRADESH<br>201310<br>India |
| **Phone** | 9739798272 |
| **Fax** | |
| **Email** | |

**Details Contact Person (Scientific Query)**

| Details Contact Person (Scientific Query) | |
|---|---|
| **Name** | Suresh Kumar Karri |
| **Designation** | Sr. Project Manager |
| **Affiliation** | Triomics Healthcare Pvt Ltd |
| **Address** | Level 5, Green Boulevard, Block C, Sector 62 Noida Uttar Pradesh 201 301 India<br>Gautam Buddha Nagar<br>UTTAR PRADESH<br>201310<br>India |
| **Phone** | 9739798272 |
| **Fax** | |
| **Email** | |

**Details Contact Person (Public Query)**

| Details Contact Person (Public Query) | |
|---|---|
| **Name** | Suresh Kumar Karri |
| **Designation** | Sr. Project Manager |
| **Affiliation** | Triomics Healthcare Pvt Ltd |
| **Address** | Level 5, Green Boulevard, Block C, Sector 62 Noida Uttar Pradesh 201 301 India<br>Gautam Buddha Nagar<br>UTTAR PRADESH<br>201310<br>India |

| | |
|---|---|
| **Phone** | 9739798272 |
| **Fax** | |
| **Email** | |

**Source of Monetary or Material Support**

| Source of Monetary or Material Support |
|---|
| > Ultrahuman Healthcare Pvt Ltd 45/3, Residency Rd, Shanthala Nagar, Ashok Nagar, Bengaluru, Karnataka 560025 |

**Primary Sponsor**

| Primary Sponsor Details | |
|---|---|
| **Name** | Ultrahuman Healthcare Pvt Ltd |
| **Address** | 1st Floor Gopala Krishna Complex, #45/3 Residency Road, Bangalore, Karnataka, India - 560025 |
| **Type of Sponsor** | Other [A preventive healthcare technology] |

**Details of Secondary Sponsor**

| Name | Address |
|---|---|
| NIL | NIL |

**Countries of Recruitment**

| List of Countries |
|---|
| India |

**Sites of Study**

| Name of Principal Investigator | Name of Site | Site Address | Phone/Fax/Email |
|---|---|---|---|
| Dr R P Rajesh | Aadhavvan Diabetes & Research Centre | No.3, 5th Street, Eswar Nagar, Kodambakkam, Chennai - 600024<br>Chennai<br>TAMIL NADU | 9629394222<br><br> |
| Dr Prabhat Ranjan Sinha | Aakash Healthcare Private Limited Hospital | Plot Road No. 201, Sector-3, Dwarka, New Delhi -110075<br>South West<br>DELHI | 9811709628<br><br> |
| Dr Neeta Deshpande | Belgaum Diabetes Centre | Ground and second floor, maruti street, Belgaum- 590001<br>Belgaum<br>KARNATAKA | 9880271313<br><br> |
| Dr Banshi Saboo | Diacare Research | 1,2 Gandhi Park, near nehrunagar, Ambawadi, Ahmedabad- 380015<br>Ahmadabad<br>GUJARAT | 8375079880<br><br> |
| Dr Suresh S M | Dr. Suresh Diacare | No 723B, 11th Main Rd, 3rd Block, Rajajinag, Bengaluru 560010<br>Bangalore<br>KARNATAKA | 9844011862<br><br> |
| Dr BVSN Raju | Induss Hospital | Opposite Kothapet Fruit Market Kothapet, Sri Sai Shivani Complex, HUDA Complex, Saroornagar 500035<br>Hyderabad<br>TELANGANA | 9440383778<br><br> |
| Dr Sriharee Kulkarni | Kulkarnis Medzone | Kulkarrni Medzone, G D Naidu Hall, Mohan Matrix,, 450, 12th Cross Rd, near Vidya Bharathi School, | 9480427359<br><br> |

| | | | |
|---|---|---|---|
| | | Mahalakshmipuram<br>Bangalore<br>KARNATAKA | |
| Dr Pankaj Aneja | Naveda Healthcare Center | A-1/81, Sector - 8, Rohini, Delhi 110085<br>North West<br>DELHI | 9811117266<br><br> |
| Dr Hamsraj Alva | Vinaya Hospital and Research Centre (a unit of KIMS) | Karangalpady, Mangaluru-575003<br>Dakshina Kannada<br>KARNATAKA | 9343562622<br><br> |

**Details of Ethics Committee**

| Name of Committee | Approval Status | Date of Approval | Is Independent Ethics Committee? |
|---|---|---|---|
| Aakash Healthcare Private Limited | Approved | 11/08/2022 | No |
| Bangalore Ethics Committee | Approved | 03/10/2022 | Yes |
| Bangalore Ethics Committee | Approved | 26/09/2022 | Yes |
| Diabetes centre ethics committee | Approved | 12/08/2022 | No |
| Ethics committee Vinaya Hospital | Approved | 30/07/2022 | No |
| Independent ethics committee | Approved | 02/08/2022 | Yes |
| Independent ethics committee - Universal Ethics Committee | Approved | 08/08/2022 | Yes |
| Induss Hospital Institutional Ethics Committee | Approved | 14/10/2022 | Yes |
| Shrey hospital Instituitional ethics committee | Approved | 12/08/2022 | No |

**Regulatory Clearance Status from DCGI**

| Status | Date |
|---|---|
| Not Applicable | No Date Specified |

**Health Condition / Problems Studied**

| Health Type | Condition |
|---|---|
| Healthy Human Volunteers | Non-diabetic and prediabetic participants. |

**Intervention / Comparator Agent**

| Type | Name | Details |
|---|---|---|

**Inclusion Criteria**

| Inclusion Criteria | |
|---|---|
| **Age From** | 25.00 Year(s) |
| **Age To** | 50.00 Year(s) |
| **Gender** | Both |
| **Details** | General inclusion criteria for healthy and prediabetic population:<br/> Subjects within the age group of 25-50 years.<br/> Subjects willing to participate in the study. <br/> BMI within 20 – 30 kg per m2<br/> Willing to comply to the advised use of CGM, activity tracker, CGM reader and the UH Application.<br/> Inclusion criteria for healthy population:<br/> Fasting blood glucose level at screening 79-99 mg/dl <br/> HbA1c range 4.0-5.6 percentage<br/> 2-hour plasma glucose during 75-g OGTT below 140 mg/dL (less than 7.8 mmol/L)<br/> Inclusion criteria for prediabetic population:<br/> Fasting blood glucose level at screening 100-125 mg/dl OR<br/> |

| | |
|---|---|
| | HbA1c range 5.7-6.4 percentage OR<br/> 2-hour plasma glucose during 75-g OGTT 140–199 mg/dL (7.8–11.0 mmol/L)<br/> |

**Exclusion Criteria**

| Exclusion Criteria | |
|---|---|
| **Details** | History of acute or subacute infection in the last three months.<br>History of chronic illnesses and autoimmune conditions.<br>Subjects on antimicrobial drug agents, including antibiotics, antivirals, or antifungals<br>Subjects pre-diagnosed with Type 1 Diabetes<br>Subjects pre-diagnosed with Type 2 Diabetes Mellitus according to the ADA (American Diabetes Association) criteria.<br>Subjects with anemia [less than male 13 grams per dL and females less than 12 grams per dL]<br>Documented known medical history of any impaired renal and liver function.<br>Subjects diagnosed with cardiac disease, including angina, heart failure, arrhythmias, valvular heart disease, or congenital heart disease.<br>Subjects with regular alcohol consumption of more than eight drinks per week and 15 drinks per week in women and men, respectively.<br>Enrolment in any kind of weight reduction program in the last six months<br>Unexplained intentional or unintentional weight loss of more than 10 percent of average weight in the previous six months<br>Subjects currently on diet plans such as keto, low carb and intermittent fasting.<br>Pregnant and lactating females<br>Out of range results in any of the screening tests carried out for blood glucose measurements (FBG, HbA1c and OGTT tests) |

| | |
|---|---|
| **Method of Generating Random Sequence** | Not Applicable |
| **Method of Concealment** | An Open list of random numbers |
| **Blinding/Masking** | Not Applicable |

**Primary Outcome**

| Outcome | Timepoints |
|---|---|
| CGM-based glucose indices over 14 days of period<br>Mean glucose levels described by a 24-hour profile during 2 weeks<br>Time in glucose ranges<br>Glycaemic variability as measured by the standard deviation, coefficient of variation, MAGE | Day 1 and Day 14 |

**Secondary Outcome**

| Outcome | Timepoints |
|---|---|
| Changes in FBS from day 0 to day 15<br>Assessing the correlations between the biomarkers and metabolic health metrics, the key relationships to explore are:<br>Glucose variability indices and sleep duration<br>Glucose variability indices and stress (as measured by cortisol)<br>Glucose variability indices and inflammation (as measured by Hs-CRP)<br>Glucose variability indices and gut microbiome index<br>Glucose variability indices and urine metabolites<br>Glucose variability indices and physical activity (limited to step count and heart rate) | Day 0 and Day 14 |

| | |
|---|---|
| **Target Sample Size** | **Total Sample Size=**100<br>**Sample Size from India=**100<br>**Final Enrollment numbers achieved (Total)=**Applicable only for Completed/Terminated trials<br>**Final Enrollment numbers achieved (India)=**Applicable only for Completed/Terminated trials |
| **Phase of Trial** | N/A |
| **Date of First Enrollment (India)** | 24/08/2022 |
| **Date of First Enrollment (Global)** | No Date Specified |
| **Estimated Duration of Trial** | **Years=**0<br>**Months=**3<br>**Days=**0 |
| **Recruitment Status of Trial (Global)** | Not Applicable |
| **Recruitment Status of Trial (India)** | Closed to Recruitment of Participants |
| **Publication Details** | None yet |
| **Brief Summary** | In this prospective observational study, eligible subjects willing to participate in the study will be screened, and the subjects meeting the eligibility criteria will be enrolled in the study. Based on the ADA criteria of Screening and Diagnostic Tests for Prediabetes, the subjects will be assigned to either group A (healthy non-diabetic subjects) or group B (pre-diabetic subjects). After the screening of the subjects, urine, stool, and blood samples will be collected for baseline investigations. All the enrolled subjects will be provided with the CGM device and trained by the on-site staff on applying it to their arms. The CGM data-collection period will be 14 days (2 weeks). Based on GV Indices, Activity & Sleep data, the UH platform will provide Nudges related to GV Indices trend. Participants can modify their diet & activities accordingly. The glucose readings will be collected by the Abbott freestyle libre CGM device, which will be scanned with the subject’s phone and the supplied Abbott CGM reader. The daily food consumption will be logged on the UH application. The activity tracker will log |

| | |
|---|---|
| | the physical activities, sleeping, and waking time. All participants will be provided with the necessary training to familiarize themselves with the App’s features and use. |
